# Rapidly Measuring Spatial Accessibility of COVID-19 Healthcare Resources: A Case Study of Illinois, USA

**DOI:** 10.1101/2020.05.06.20093534

**Authors:** Jeon-Young Kang, Alexander Michels, Fangzheng Lyu, Shaohua Wang, Nelson Agbodo, Vincent L Freeman, Shaowen Wang

## Abstract

The severe acute respiratory syndrome coronavirus 2 (SARS-CoV-2), causing the coronavirus disease 2019 (COVID-19) pandemic, has infected millions of people and caused hundreds of thousands of deaths. While COVID-19 has overwhelmed healthcare resources (e.g., healthcare personnel, testing resources, hospital beds, and ventilators) in a number of countries, limited research has been conducted to understand spatial accessibility of such resources. This study fills this gap by rapidly measuring the spatial accessibility of COVID-19 healthcare resources with a particular focus on Illinois, USA. Specifically, the rapid measurement is achieved by resolving computational intensity of an enhanced two-step floating catchment area (E2SFCA) method through a parallel computing strategy based on cyberGIS (cyber geographic information science and systems). The study compared the spatial accessibility measures for COVID-19 patients to those of general population, identifying which geographic areas need additional healthcare resources to improve access. The results also help delineate the areas that may face a COVID-19-induced shortage of healthcare resources caused by COVID-19. The Chicagoland, particularly the southern Chicago, shows an additional need for resources. Our findings are relevant for policymakers and public health practitioners to allocate existing healthcare resources or distribute new resources for maximum access to health services.

## 1. Introduction

A novel coronavirus disease (named COVID-19) caused by severe acute respiratory syndrome coronavirus 2 (SARS-CoV-2) has widely spread worldwide. As of May 6, 2020, about 3.6 million COVID-19 cases have been confirmed worldwide; and in the United States alone, over 1.2 million people have been sick with more than 73,000 deaths. Among the infected cases, hundreds of thousands of people are hospitalized. The COVID-19 pandemic has exceeded the capacities of healthcare resources, including healthcare personnel, testing resources, hospital beds, and ventilators (White and Lo, 2020; Xie et al., 2020). This increased demand for healthcare resources has caused or exacerbated disparities in access to healthcare.

Oftentimes, COVID-19 infection cases and related healthcare resources in demand are not spatially proportionally distributed. In other words, spatial mismatches need to be resolved between healthcare resource availability and the needs of COVID-19 patients and general population. Identifying these mismatches is crucial for allocating healthcare resources efficiently and effectively (McLafferty, 2015). Therefore, understanding to what extent the healthcare resources are available in a particular spatial context is urgent.

Measuring spatial accessibility to healthcare resources has long been of interest to public health research. Examples include healthcare access for seniors (Gu et al., 2009; Paez et al., 2010), disabled people (Church and Marston, 2003), cancer-specific survivals (Freeman et al., 2020; Wan et al., 2012), examining spatial accessibility among populations with multiple transportation modes (Mao and Nekorchuk, 2013; Kaur Khakh et al., 2019), and access to specific health care treatments such as mammograms (Henry et al., 2013). Spatial accessibility of healthcare resources can be measured by spatial interactions between the amount of supplies (e.g., the number of hospital beds or physicians) and demands along with the distance and travel time between the locations of healthcare resources and those of residential areas.

A commonly used method for measuring spatial accessibility is the two-step floating catchment area (2SFCA) method (Luo and Wang, 2003). Particularly, the enhanced two-step floating catchment area (E2SFCA) method accounts for distance decay (Luo and Qi, 2009). The E2SFCA method uses travel time for a given mode of transportation to derive the areas that are within 10, 20, and 30 minutes of supply locations. For large study areas such as the state of Illinois in the USA, calculating catchment areas on a road network has high computational intensity (Wang and Armstrong, 2009). CyberGIS – defined as geographic information science and systems based on advanced cyberinfrastructure – is well suited to resolve this type of computational intensity through high-performance parallel computing (Kang et al., 2020; Wang, 2010; Wang, 2016).

Motivated to address this computationally intensity challenge, this study aims to rapidly measure the spatial accessibility of healthcare resources in Illinois. Specifically, the study seeks to answer the following two research questions: (1) to what extent Illinois residents have access to healthcare resources during the COVID-19 pandemic and (2) which geographic areas have resource surplus and which areas have resource shortage? To answer these questions, spatial accessibility was measured based on travel time between the locations of residence and healthcare resources in the context of COVID-19 patients and general population.

We have developed a parallel enhanced two-step floating catchment area (P-E2SFCA) method based on CyberGIS-Jupyter – a cyberGIS framework for achieving data-intensive, reproducible, and scalable geospatial analytics using Jupyter Notebook (Kang et al., 2019; Lyu et al., 2019; Wang et al., 2020; Yin et al., 2019). In our study, the demand of healthcare resources comes from either COVID-19 patients or general population. Here, the general population is considered in the analysis to assess the generalized need for healthcare resources during an uncontrolled outbreak. We compare spatial accessibility of healthcare between COVID-19 patients and the general population. Our analysis provides insights for decision making to optimize the allocation of COVID-19 related healthcare resources, such as hospital beds, testing resources, healthcare personnel, and ventilators. The following section (Section 2) describes the data and method used in the study. Section 3 summarizes and evaluates the results obtained from the P-E2SFCA method. Section 4 concludes with a discussion about policy implications and future directions.

## 2. Data and Method

### 2.1. Study Area and Data

This study focuses on spatial accessibility of healthcare resources for the general population and COVID-19 patients in Illinois, USA. We used four types of datasets, summarized as follows: 1) hospital dataset, 2) COVID-19 confirmed case dataset, 3) residential dataset, and 4) road network dataset. The hospital dataset was provided by the Homeland Infrastructure Foundation-Level Data (HIFLD) open data forum (https://hifld-geoplatform.opendata.arcgis.com/). The HIFLD provides national-level geospatial data that can be used in supporting community preparedness and research. The COVID-19 confirmed case dataset at the zip-code level is provided by the Illinois Department of Public Health (IDPH) (http://dph.illinois.gov/covid19/covid19-statistics). The residential dataset was obtained from the United States Census Bureau. We used an Application Programming Interface (API) call for pulling data from the 2018 American Community Survey 5-year detail table for the Zip Code Tabulation Areas (ZCTAs) in the state of Illinois, USA. The road network dataset was retrieved using a Python package OSMnx (Boeing, 2017), which helped us to download and analyze street networks from the OpenStreetMap.

In Illinois, there are 183 hospitals from which the information of beds is available in the dataset. We excluded some hospitals, including military, children, psychiatric, and rehabilitation in this study, because these types of hospitals may not provide health services to COVID-19 patients. Figure 1(a) illustrates the spatial distribution of hospitals with the population across Illinois. Figure 1(b) shows the spatial distribution of the COVID-19 confirmed cases in Illinois as of April 10, 2020. Obviously, the population in Illinois is dense in the Chicago area, and significant clustering of the COVID-19 cases have occurred in Chicago. As of April 10, 2020, about 18,000 COVID-19 cases were confirmed out of about 88,000 tests. In Chicago, there are 29 hospitals in which beds are available for the patients and the general population (Figure 2(a)). As of April 10, 2020, there were about 7,600 confirmed COVID-19 cases (Figure 2(b)).

**Figure 1.**
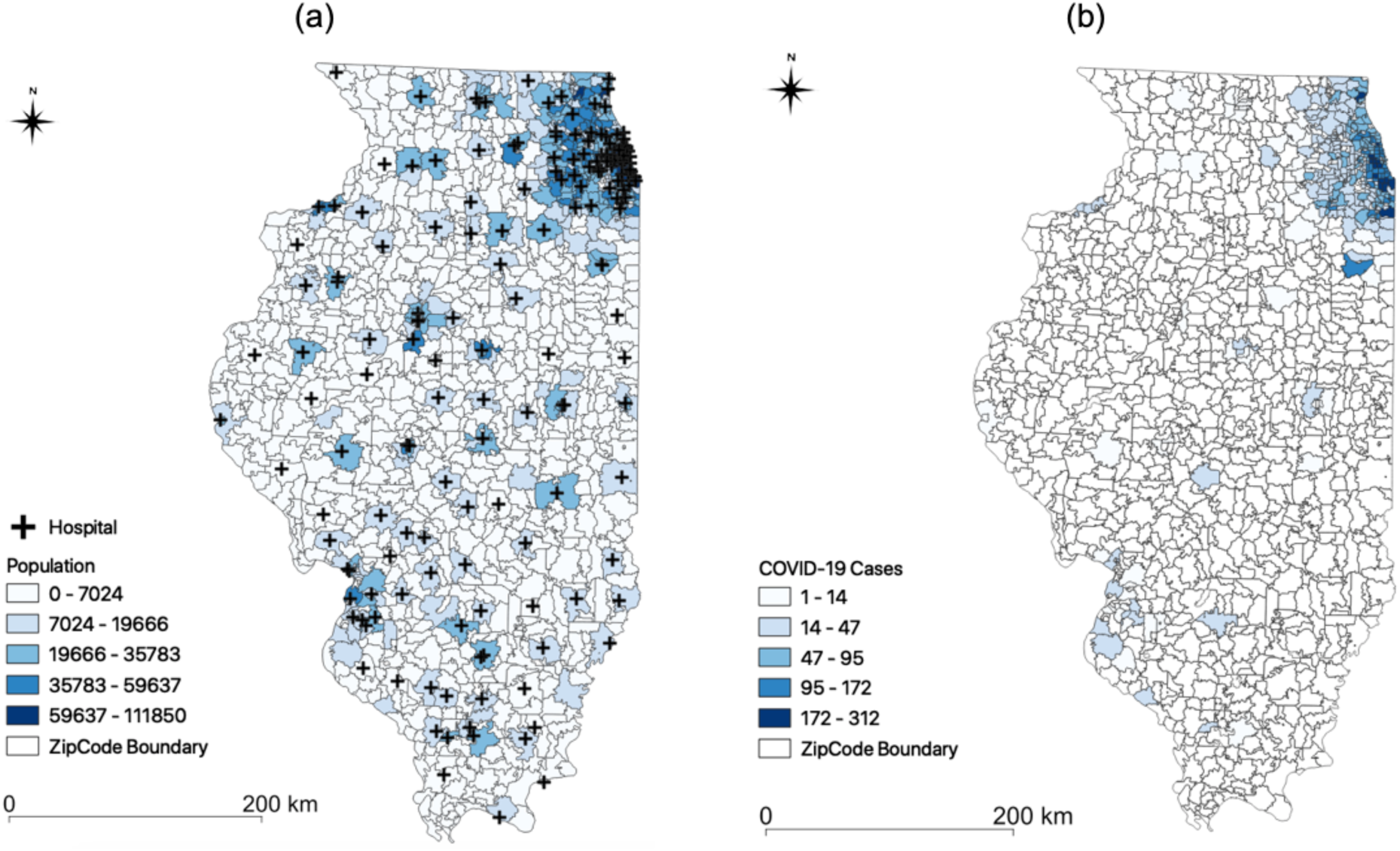
Datasets of hospital, population, COVID-19 cases in Illinois: (a) spatial distribution of hospitals and population; (b) spatial distribution of confirmed COVID-19 cases.

**Figure 2.**
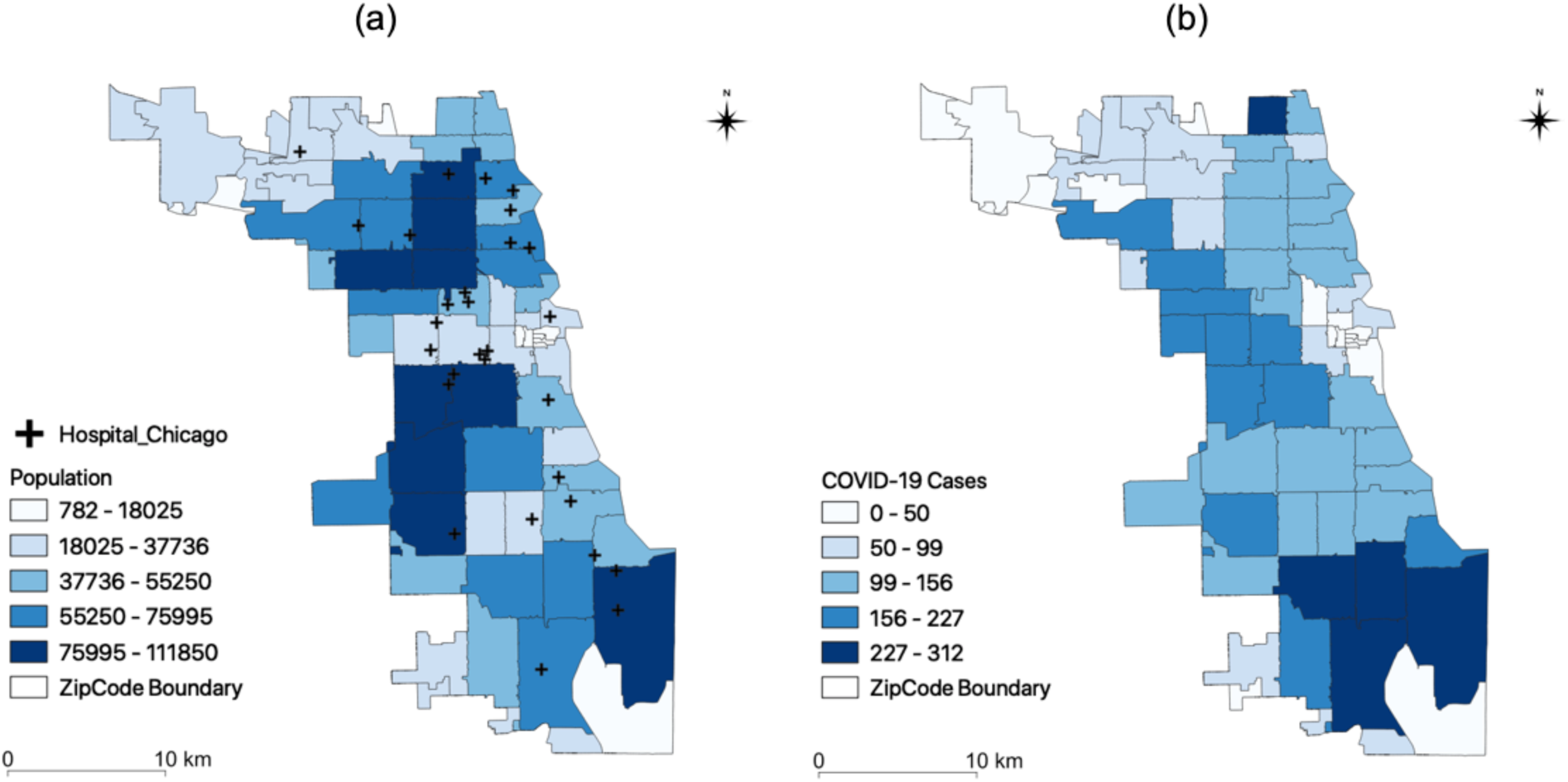
Datasets of hospitals, population, COVID-19 cases in Chicago, Illinois: (a) spatial distribution of hospitals and population, and (b) spatial distribution of confirmed COVID-19 cases.

### 2.2. Parallel Enhanced Two-Step Floating Catchment Area (P-E2SFCA) Method

The conventional two-step floating catchment area (2SFCA) method is based on a service-to-population ratio computed in two steps (Luo and Wang, 2003; Guagliardo 2004). The first step consists of finding all people (*i*) located within a catchment area of each healthcare (*j*), as shown in Figure 3(a). A catchment area is based on a threshold travel time (*d*_0_). The second step computes a service-to-population rate *R_j_* within a catchment area.

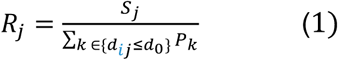

where for the number of available resources*(s)* at each healthcare (*j*), find all population locations (*k*) that fall within a threshold travel distance (*d*_0_).

**Figure 3.**
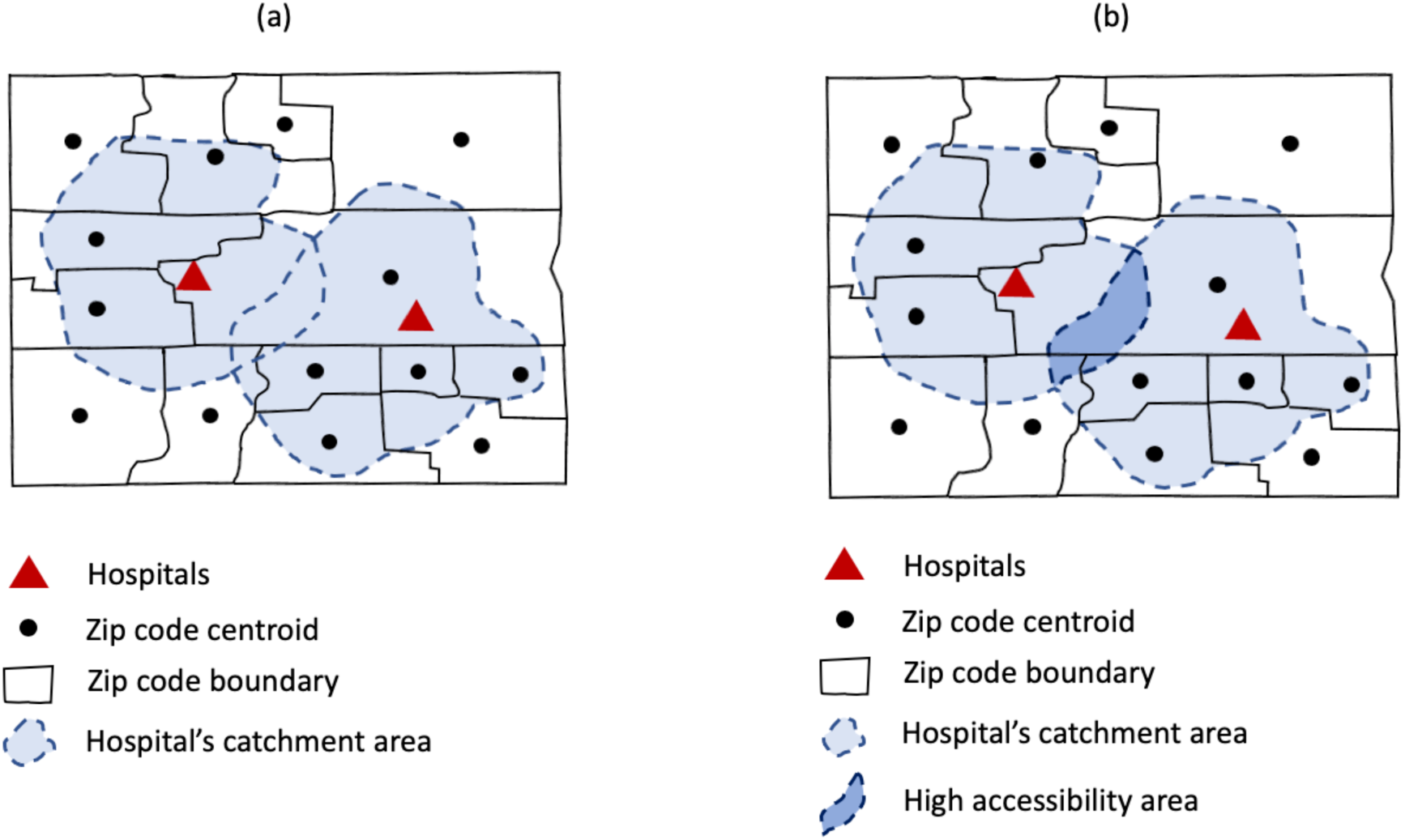
Measure of spatial accessibility of a set of hospitals: (a) delineate the catchment area of each hospital; and (b) sum up the accessibility value at residential locations.

Then, the accessibility *A_i_* at a residential location *i* is computed by summing up the service-to-population ratios, as shown in Figure 3 (b).

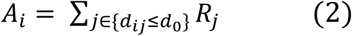

where *i* denotes a residential location, and *R_j_* is the proportion of services per person at healthcare location *j* whose centroids are located within the catchment area.

As a relative measurement, the accessibility measurement *A_i_* indicates which geographic areas are relatively more accessible than other areas. For example, overlapped areas in Figure 3(b) may have a relatively higher accessibility than other areas. The areas that do not fall within catchment areas of any hospitals have no access to any hospitals within a predetermined travel time (e.g., 30 minutes).

To fully take into account the residents who may be more likely to visit closer hospitals than others, an enhanced two-step floating catchment area (E2SFCA) method was developed (Luo and Qi, 2009) to resolve the limitation about no distance decay within a catchment area (i.e., residents in the same catchment area are assumed to have equal spatial accessibility). The E2SFCA method accounts for the distance decay, by allowing for multiple travel time zones, such as 0-10, 10-20, and 20-30 minutes. Three values of weights (1, 0.68, and 0.22) were applied to each travel time zone (0-10, 10-20, and 20-30 mins), respectively (Luo and Qi, 2009). Spatial accessibility is therefore computed as a summation of the measure of accessibility at each travel interval.

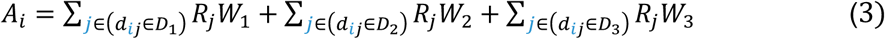

where *A_i_* denotes the accessibility of people at location *i* to hospitals, and the proportions of service-to-population *R_j_* at healthcare locations *j* whose centroids are located within the catchment area. *W_r_* denotes the distance weight for *rth* travel intervals.

Figure 4 (a) shows the workflow of the E2SFCA method using hospital beds as a case of healthcare resources. The method has two major steps: calculating a bed-to-population ratio for each supply location and then summing these ratios for residential locations where supply regions overlap. Since we aim to measure the accessibility for the general population and COVID-19 patients, we separated the calculation of a bed-to-population ratio from that of a bed-to-COVID-19-patients ratio. The first step delineates every hospital’s 30-minute driving zone through a convex hull. Each zone is segmented into 0-10, 10-20, and 20-30 minutes sub driving zones. These zones are used to compute a hospital bed-to-population ratio using a weighted sum of residential locations within each hospital’s catchment area following equation (3). Lastly, the accessibility measurements are aggregated into hexagon grids to minimize orientation bias from edge effects (Figure 5).

**Figure 4.**
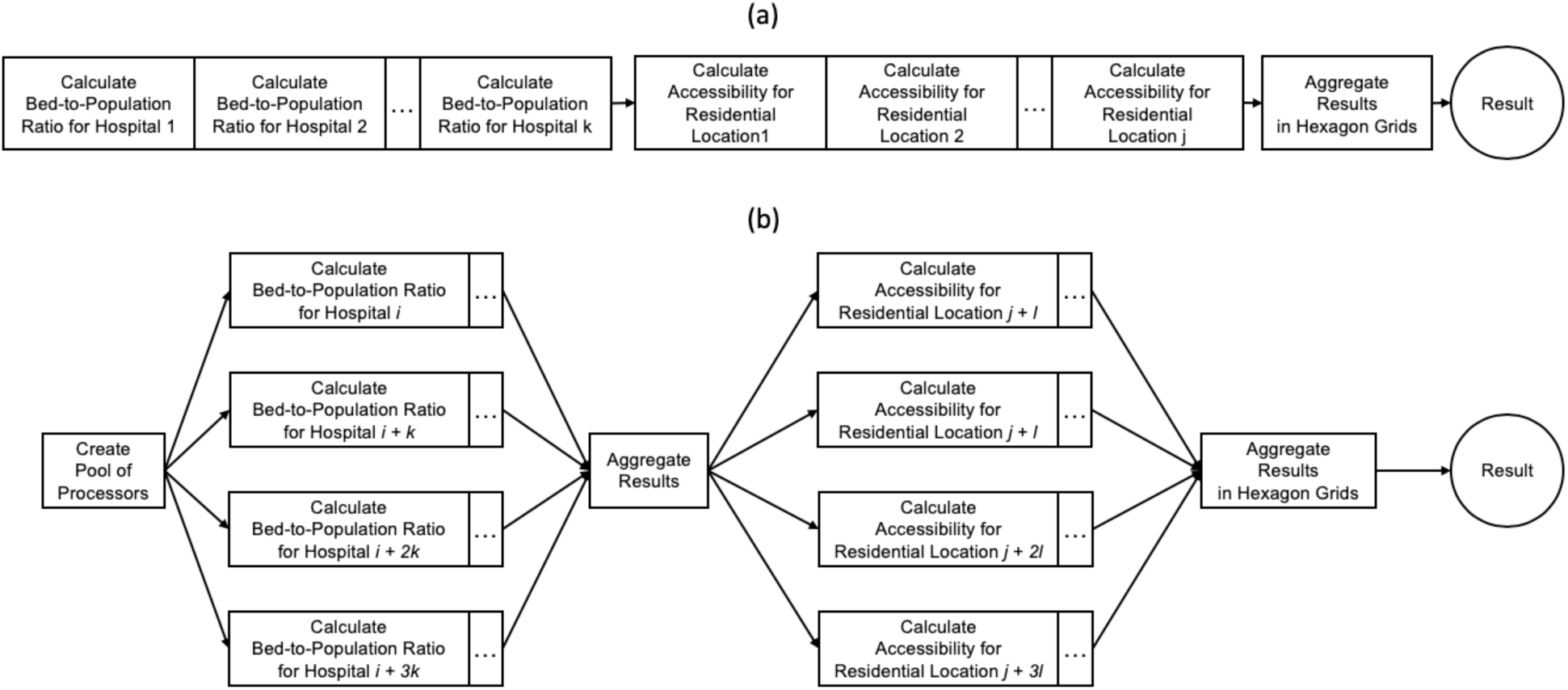
Computational workflows: (a) E2SFCAM; and (b) P-E2SFCAM

**Figure 5.**
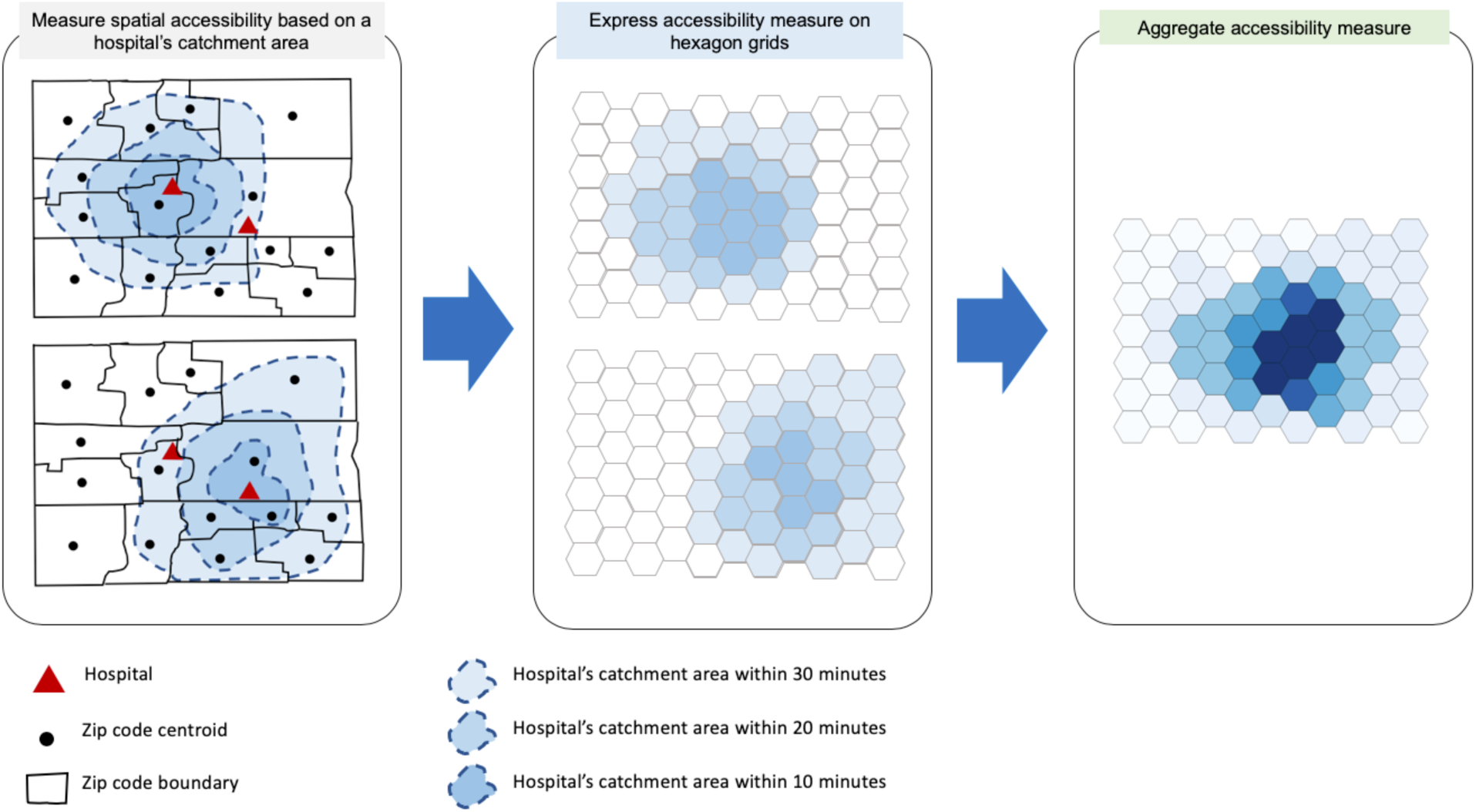
Expression of accessibility measures on hexagon grids. Note: Darker colors represent areas with higher accessibility. The values of hexagons are determined by *A_i_* from equation (2).

Each hospital’s catchment area and its corresponding driving zones can be expressed on hexagon grids (Figure 5). If there are overlapping catchment areas, the values at each hexagon grid are aggregated. Specifically, we used 500-meter hexagon grids for Chicago and 5-km hexagon grids for Illinois. In Chicago, about 400 meters are the shortest distance between zip code centroids. Therefore, 500-meter hexagon grids are sufficient to represent the accessibility measure. Considering the relatively large area of Illinois, 5-km hexagon grids are suitable for depicting the accessibility measure.

Calculating this measure is computationally intensive because creating driving zones generally relies on defining a boundary of nodes within a road network that are at the edge of the driving zones, generating a convex hull around these nodes, and then querying if residences lie within the boundary. The method also depends on calculating intersections and differences between multiple hospitals’ driving zones and aggregating the collected statistics appropriately for each population center.

In order to obtain analytical results rapidly, which is important for the fight against the COVID-19 crisis, we have developed a parallel computing approach to speeding up the E2SFCA method. We parallelized the steps of calculating an egocentric graph of the road network within specified travel times, determining the convex hull of the nodes, and calculating the difference between convex hulls to derive 0-10, 10-20, and 20-30 minutes driving zones. Once these steps are completed, we also parallelized the portion of the steps that calculate intersections between the driving zones to aggregate statistics across different hospitals into a holistic measure of spatial accessibility in hexagon grids. An illustrative example of P-E2SFCA with four processors is provided in Figure 4 (b).

As a result, accessibility measures are derived. However, the absolute values of the accessibility measures may not be important to revealing which areas have lower accessibility than other areas. Instead of using the absolute value, we converted the absolute values of accessibility measure to the normalized measure as follows:

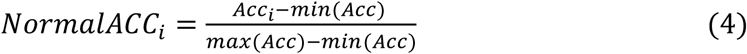

where *Acc_i_* refers to the accessibility measure at residential location *i*.

To see which areas have relatively more resources given the COVID-19 confirmed cases in Illinois, we used an existing method to measure the percentage difference in model estimation (Downs et al., 2018; Mao & Nekorchuk, 2013). The spatial accessibility for COVID-19 patients was compared to the accessibility for the general population by calculating the percentage difference, as:

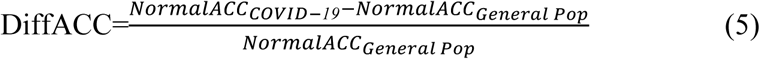

A positive value of the comparison is derived if the normalized accessibility for the COVID-19 patients is larger than that for general population, indicating the proportion of oversupply of beds. The zero value indicates that the accessibility for the COVID-19 patients and that for the general population are identical. Otherwise, if the value is negative, then the patients might have struggles to access healthcare resources. Because the values of the comparison are relative measurements, they do not directly quantify the magnitude of issues for particular areas to access healthcare resources.

## 3. Results

We used the P-E2SFCA method to assess the spatial accessibility of hospital beds for the general population and the COVID-19 patients in Chicago and Illinois, USA, as of April 10, 2020. In addition, the results from the P-E2SFCA method identified the areas in which hospital beds are relatively more or less accessible. Our analysis also helps identify the areas in which there are imbalances between the accessibility of healthcare resources and needs for the general population and the COVID-19 patients.

Based on the accessibility measure in Chicago (Figure 6), we found that the accessibility of hospital beds is spatially varying. The areas with spatial accessibility measures at or above the median are located near the center of Chicago and extending northwestward. The areas with lower accessibility measures are located in southern Chicago. In other words, people living in central and northern Chicago have better accessibility to hospital beds than those living in southern Chicago, as hospitals and hospital beds are mostly located in central Chicago.

**Figure 6.**
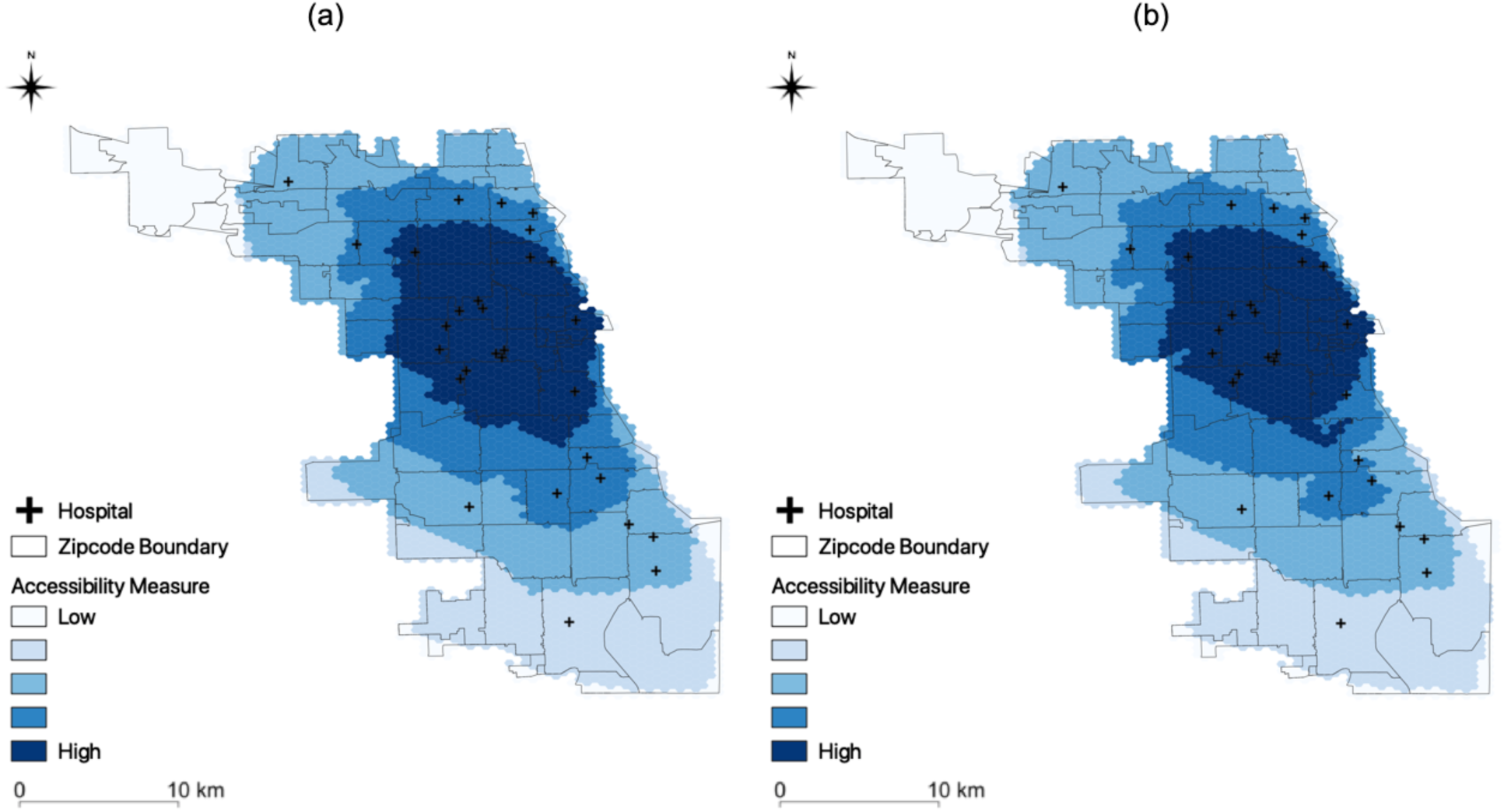
Spatial accessibility measure for Chicago: (a) general population; and (b) COVID-19 patients. The area of northwestern Chicago was excluded from analysis because that is where the O’Hare International airport is located.

In addition, the difference in access to hospital beds for the general population and for the COVID-19 patients is not statistically significant (*t*_(6556)_=-0.8185, *p*>0.05), as shown in Figure 7. This implies that access to hospital beds was similar for the general population at risk and for the patients diagnosed with COVID-19. The median of the normalized accessibility measure is equal to 0.5044 for the general population and 0.4915 for the COVID-19 patients, respectively. In general, hospital beds are distributed with high accessibility in Chicago.

**Figure 7.**
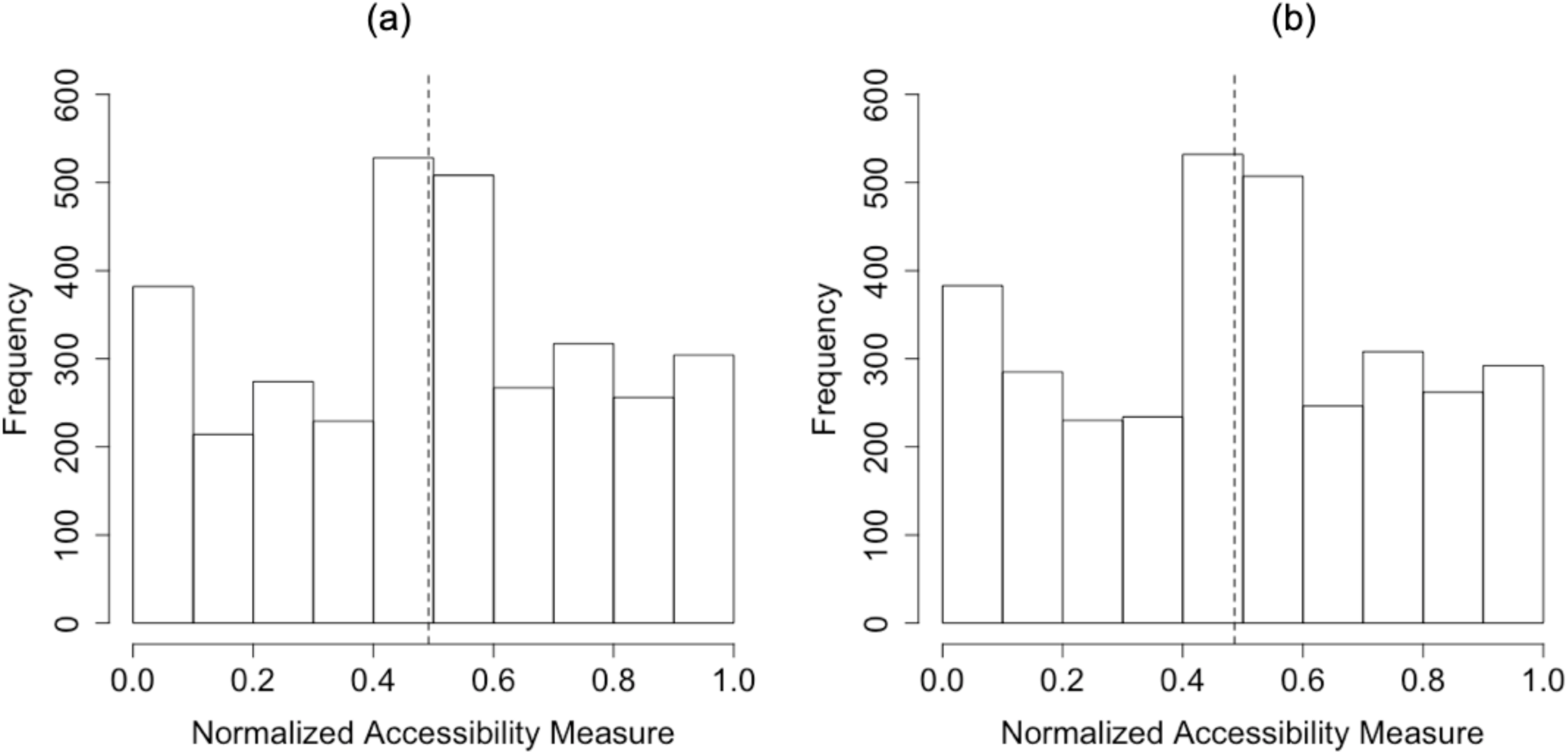
Accessibility measure for Chicago: (a) the general population (b) COVID-19 patients in the Chicago area. Note: the dotted lines refer to the average of the accessibility measure. Y-axis denotes the number of hexagons, since the accessibility measure is represented through the hexagons.

As shown in Figure 8, the spatial accessibility measure is also spatially varying across Illinois. For the general population, the areas with higher spatial accessibility are relatively uniformly distributed throughout Illinois (Figure 8(a)). On the other hand, only some areas have relatively higher spatial accessibility measures for COVID-19 patients in Illinois (Figure 8(b)). In central Illinois (e.g., Peoria and Springfield), COVID-19 patients have higher accessibility to hospital beds than those living in the other areas of Illinois. We also found that COVID-19 patients have lower accessibility in some areas of Chicago (i.e., northeastern Illinois). While abundant resources are concentrated in Chicago (i.e., northeastern Illinois), the majority of the Illinois COVID-19 cases occurred in Chicago.

**Figure 8.**
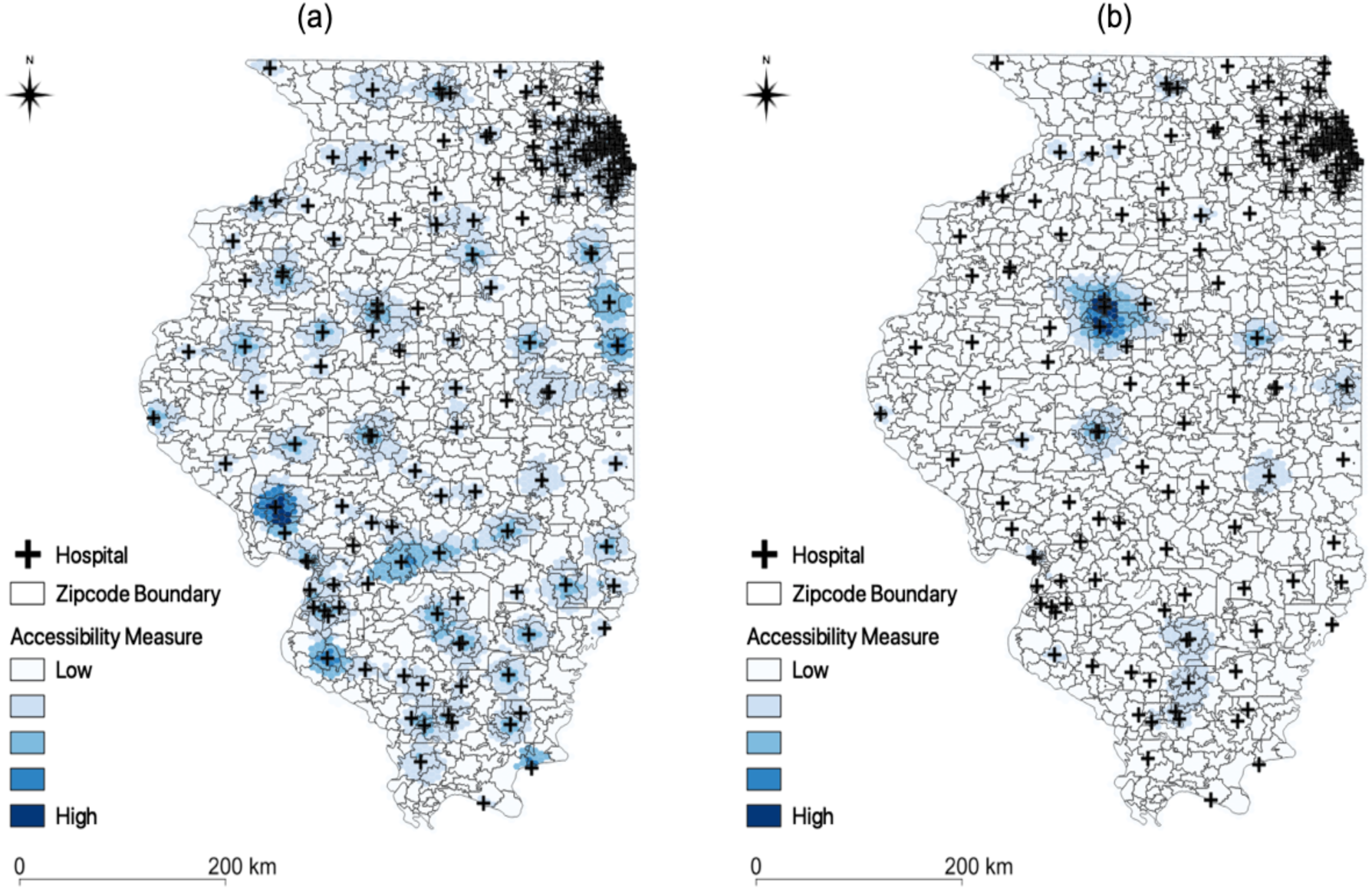
Accessibility measure for Illinois: (a) general population; (b) COVID-19 patients.

In Illinois, many residents may not have adequate access to hospital beds (Figure 9). This means that some may be struggling to get access to hospital beds at the state level. Population in urbanized areas (e.g., Chicago, Peoria, Springfield) tend to have higher accessibility to the beds. The median of the normalized accessibility measure is equal to 0.0190 for the general population and 0.0003 for the COVID-19 patients, respectively. The difference in access to hospital beds for the general population and for the COVID-19 patients is statistically significant (*t*_(13909)_=-20.086, *p*<0.001).

**Figure 9.**
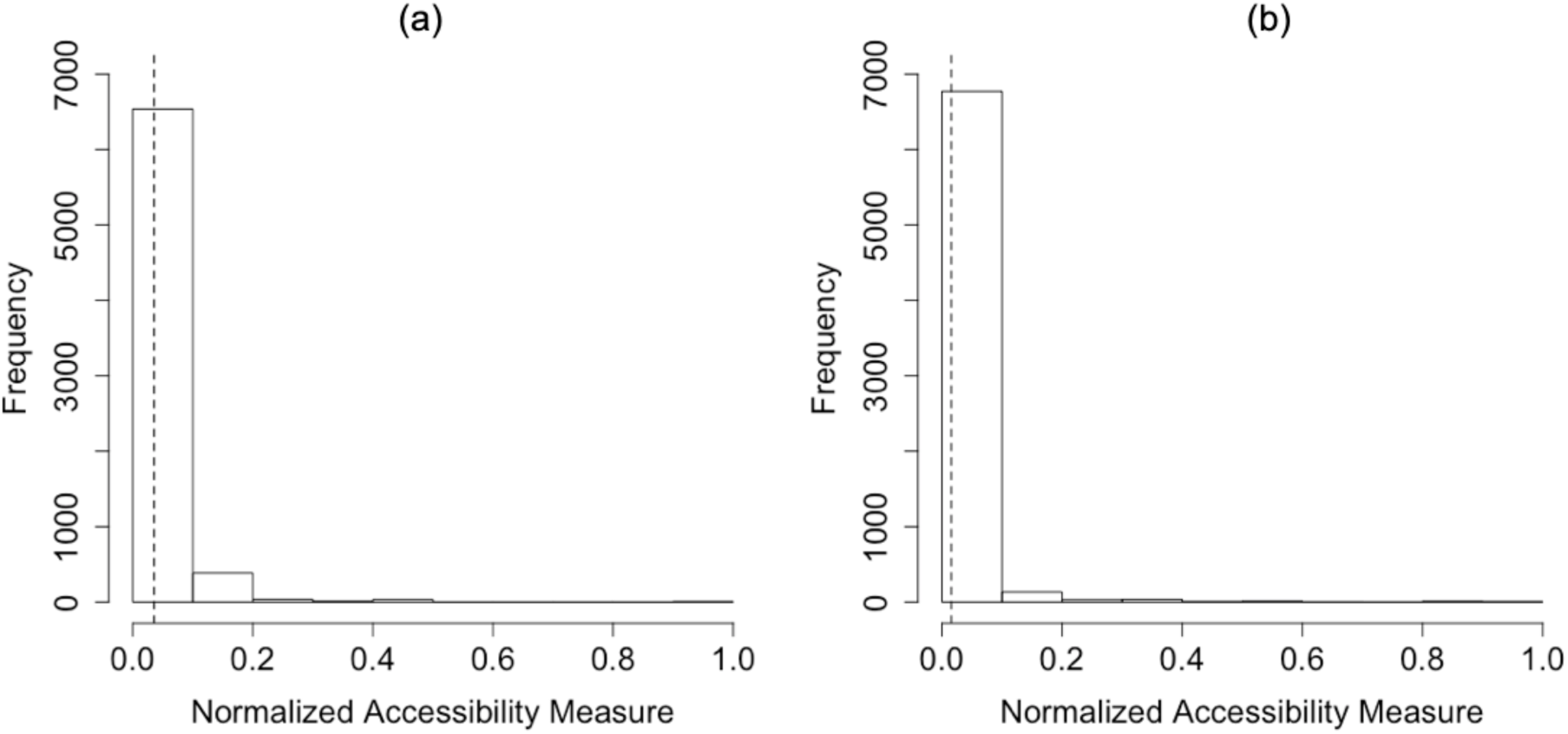
Accessibility measure for Illinois: (a) the general population; and (b) COVID-19 patients in Illinois. Note: the dotted lines refer to the average of accessibility measure. Y-axis denotes the number of hexagons, since the accessibility is represented through the hexagons.

Figure 10 presents the scatter plots elucidating whether there are imbalances between the spatial accessibility for the general population and that for COVID-19 patients in Chicago (a) and Illinois (b). Each dot represents the accessibility measure at each hexagon. Spearman’s correlation coefficients are 0.9999 (p-value <0.001) and 0.4279 (p-value <0.001) for Chicago and Illinois, respectively. If the correlation coefficient is close to 1, then there are less imbalances in the spatial accessibility between the general population and COVID-19 patients. Although the accessibility measures are not significantly different in Chicago, they are quite different in Illinois. This means that some areas may need additional beds while the beds may be abundant in other areas. In Figure 10(b), the areas having higher accessibility for the general population, but lower accessibility for COVID-19 patients could be understood as the areas in which additional beds may be needed. Furthermore, the number of COVID-19 cases is relatively higher compared to existing accessible beds in these areas. On the other hand, the areas having higher accessibility for COVID-19 patients, but lower accessibility for the general population could be defined as the areas in which hospital beds may be abundant.

**Figure 10.**
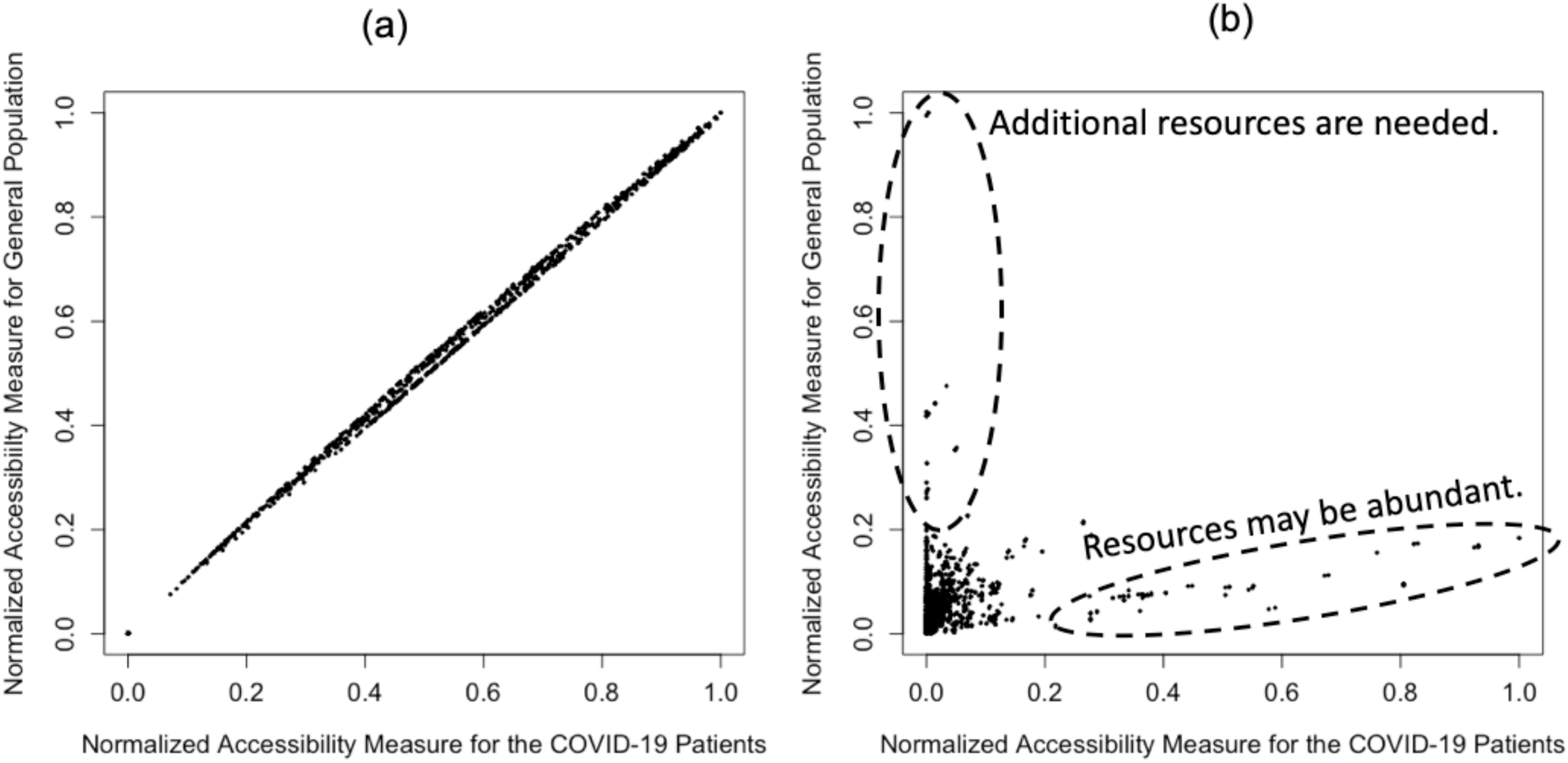
Accessibility measure: (a) Chicago; (b) Illinois.

To identify the areas that need additional beds to resolve the inequality in access to hospital beds for COVID-19 patients in Illinois, we compared the accessibility measure for COVID-19 patients to that for the general population, based on equation (5). Figure 11 illustrates the measure difference in Chicago (a) and in Illinois (b). The circles identify the areas where beds may be abundant. Compared to the accessibility for the general population, COVID-19 patients have higher accessibility in northern Chicago. In other words, it is relatively more difficult for COVID-19 patients living in southern Chicago to access hospital beds. This also implies that the hospital beds are more densely located in northern Chicago. Therefore, it is important to consider increasing the supply of beds on the southside via field hospitals and/or reopening recently closed facilities.

**Figure 11.**
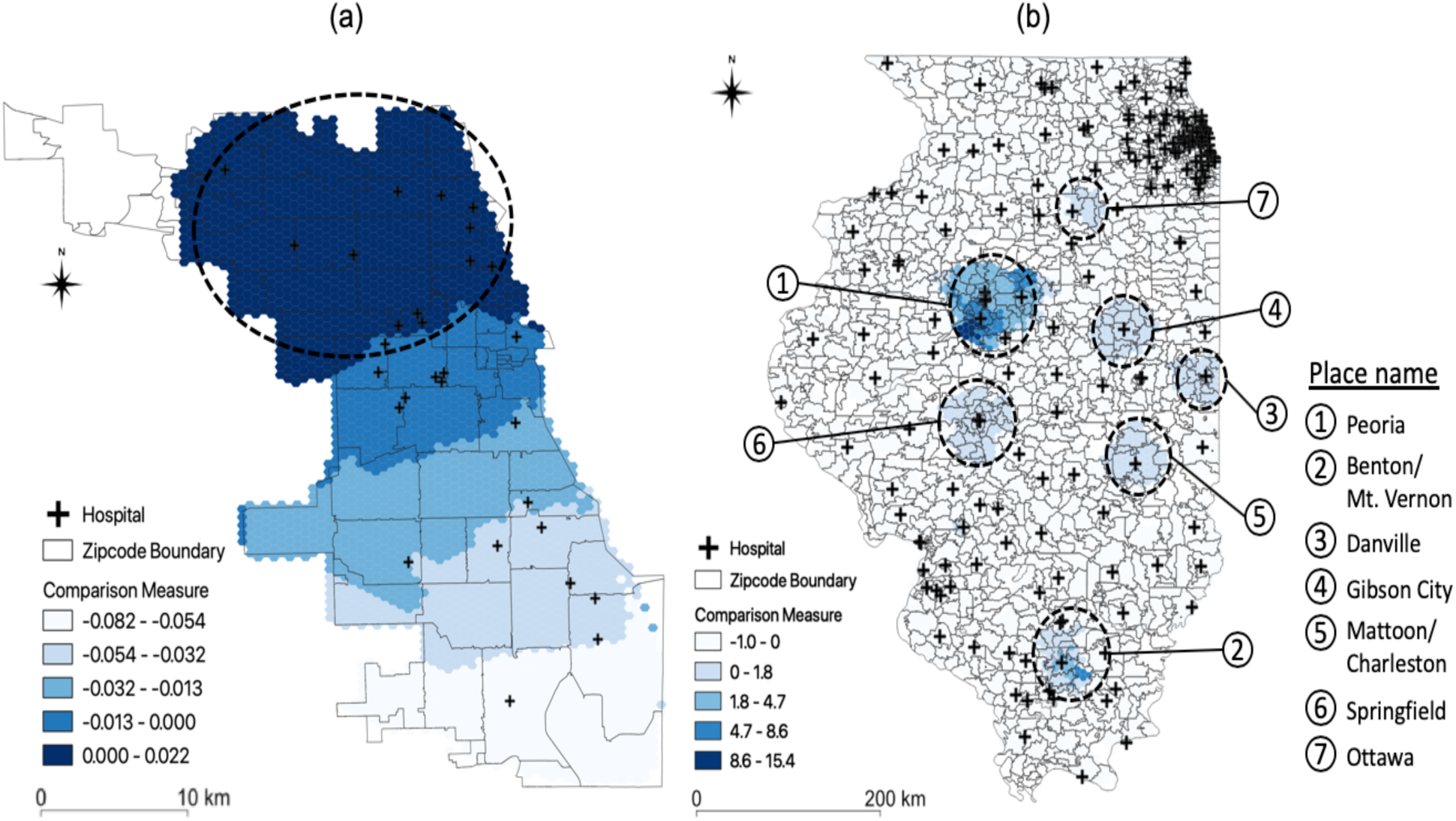
Comparison of spatial accessibility measure: (a) Chicago; and (b) Illinois.

At the state level, the hospital beds are not proportionally distributed, especially considering the number of COVID-19 confirmed cases. In Figure 11(b), the circles represent the areas in which the beds may be abundant. Peoria is the most accessible area for hospital beds, compared to the number of COVID-19 patients, and followed by Benton/Mt. Vernon, Danville, Gibson City, Mattoon/Charleston, Springfield, and Ottawa. We found that additional beds are needed in the Chicago area despite the substantial number of beds that are already available. Given that COVID-19 cases mostly occurred in Chicago, additional beds may need to be allocated to Chicago, especially the southern Chicago area.

### 3.2. Computational Performance

We assessed the computational performance of P-E2SFCA using the runtime in seconds of both P-E2SFCA and E2SFCA implementations and ran both sets of code 10 times each to calculate an average runtime in seconds. The computational experiments were conducted using CyberGIS-Jupyter and Virtual ROGER (https://cybergis.illinois.edu/infrastructures/) (Wang, 2017). For computing the accessibility in Chicago, P-E2SFCA executed at 6 times faster (i.e., an average of 942 seconds across ten runs to an average of 147 seconds) using 4 computing cores not only by parallelizing computationally intensive parts of E2SFCA, but also by efficiently aggregating results returned from parallel computing resources.

## 4. Concluding Discussion

Although extensive studies have focused on forecast (Roosa et al., 2020) and exploration of space-time patterns and trends of COVID-19 cases (Desjardins et al., 2020; Grasselli et al., 2020), research questions about the availability and capacity of healthcare resources (e.g., hospital beds, ventilators, healthcare personnel, and testing resources) for treating COVID-19 patients need rigorous investigations. This study has addressed the question of to what extent the general population and COVID-19 patients in Illinois, USA have accessibility to healthcare resources. Specifically, we compared the spatial accessibility for the general population to that for the COVID-19 patients. The comparison identifies which geographic areas may need additional healthcare resources to accommodate COVID-19 patients.

Specifically, our findings based on the P-E2SFCA method provide an improved understanding of spatial accessibility of hospital beds for the general population and the COVID-19 patients in Chicago and Illinois, USA, as of April 10, 2020. In addition, our analysis helps identify the areas in which there are imbalances between the accessibility for the general population and that for COVID-19 patients. The results also reveal which areas have relatively higher spatial accessibility of hospital beds.

Comparing the accessibility measures for the general population to that for COVID-19 patients, we found the areas that need additional healthcare resources to improve spatial accessibility in Illinois. In general, there are no significant difference in spatial accessibility measures between the general population and COVID-19 patients in Chicago. Nonetheless, given that confirmed COVID-19 cases in southern Chicago may exceed the capacity of healthcare resources available there, additional resources should be allocated to southern Chicago. At the state level, some areas (e.g., Peoria) have much higher accessibility for COVID-19 patients than for the general population. Compared to the number of confirmed cases, these areas have a resource surplus. On the other hand, the Chicago area needs more resources than other areas, considering the increasing number of confirmed cases in Chicago.

This study has several limitations. The accessibility measures near state boundaries might be underestimated because we did not include the hospitals in neighboring states (e.g., Indiana, Wisconsin, Iowa, Kentucky, and Missouri), which Illinois residents might visit. Because there are different population distribution patterns, the catchment size parameter needs to be chosen to account for the difference of such patterns (McGrail, 2012). Therefore, assessing the impact of varying catchment size on the spatial accessibility measure (Luo and Whippo, 2012) would be worth for future research. In addition, the COVID-19 cases may not be exhaustively counted because many cases were not confirmed due to having no or light symptoms (Bai et al., 2020). Also, our analysis did not distinguish between non-acute inpatient bed versus intensive care unit bed capacities. Given that the number of confirmed cases is expected to increase in Illinois, to assess spatial accessibility for COVID-19 patients over time is an important research topic. Then it would be interesting to explore the dynamics between the demand of COVID-19 patients and the supply of healthcare resources. Although our results may help identify the areas where resources are abundant or insufficient, we did not assess how much resources need to be allocated, which should be another future research topic. While our cyberGIS approach to parallel computing of E2SFCA achieves significant computational performance gains, future work needs to be done to determine the most optimal means to resolving the computational intensity, especially for the purpose of applying the analysis to larger geographic areas or to national and international assessment.

In summary, rapidly measuring spatial accessibility of healthcare resources is critical to the fight against the COVID-19 crisis, particularly for better understanding how well the healthcare infrastructure is equipped to save people’s lives. Based on this study, policymakers and public-health officials should consider optimizing the allocation of existing healthcare resources or putting additional resources into low accessibility areas. At the same time, strict quarantine, social distancing, and isolation of known cases by individuals and communities are important to slow down the spread of COVID-19 (Anderson et al., 2020; Wilder-Smith & Freedman, 2020), which in turn, help to address the important spatial accessibility issues.

## Data Availability

The data used in this study were retrieved from publicly available sources.

https://hifld-geoplatform.opendata.arcgis.com/

## Acknowledgements

This article and associated materials are based in part upon work supported by the National Science Foundation (NSF) under grant numbers: 1443080 and 1743184. Any opinions, findings, and conclusions or recommendations expressed in this material are those of the authors and do not necessarily reflect the views of NSF. Our computational work used Virtual ROGER, which is a cyberGIS supercomputer supported by the CyberGIS center for Advanced Digital and Spatial Studies and the School of Earth, Society and Environment at the University of Illinois at Urbana-Champaign.

